# Rapid Screening for SARS-CoV-2 Variants of Concern in Clinical and Environmental Samples Using Nested RT-PCR Assays Targeting Key Mutations of the Spike Protein

**DOI:** 10.1101/2021.03.09.21252450

**Authors:** G. La Rosa, P. Mancini, G. Bonanno Ferraro, C. Veneri, M. Iaconelli, L. Lucentini, L. Bonadonna, S. Brusaferro, D. Brandtner, A. Fasanella, L. Pace, A. Parisi, D. Galante, E. Suffredini

## Abstract

New SARS-CoV-2 mutations are constantly emerging, raising concerns of increased transmissibility, virulence or escape from host immune response.

We describe a nested RT-PCR assay (∼1500 bps) to detect multiple key spike protein mutations distinctive of the major known circulating SARS-CoV-2 variants, including the three Variants of Concern (VOCs) 20I/501Y.V1 (United Kingdom), 20H/501Y.V2 (South Africa), and 20J/501Y.V3 (Brazil), as well as the 20E.EU1 variant (Spain), the CAL.20C recently identified in California, and the mink-associated variant (GR, lineage B.1.1.298). Prior to application to field samples, the discriminatory potential of this PCR assay was explored using GISAID and Nextclade. To extend variant detection to challenging matrices such as sewage, where the amplification of long fragments is problematic, two short nested RT-PCR assays (∼300 bps) were also designed, targeting portions of the region spanned by the long nested assay.

The three newly-designed assays were then tested on field samples, including 7 fully-sequenced viral isolates from swab samples and 34 urban wastewater samples, some of which collected in areas where circulation of VOCs had been reported.

The long assay successfully amplified all the previously characterized viral isolates, allowing the correct identification of variants 20I/501Y.V1 and 20E.EU1 present in the panel. The sequences obtained using the short assays were consistent with those obtained with the long assay. Mutations characteristic of VOCs (UK and Brazilian variant) and of other variant (Spanish) were detected in sewage samples. To our knowledge, this is the first evidence of the presence of sequences harboring key mutations of 20I/501Y.V1 and 20J/501Y.V3 in urban wastewaters, highlighting the potential contribution of wastewater surveillance to explore SARS-CoV-2 diversity.

The developed nested RT-PCR assays can be used as an initial rapid screening test to select clinical samples containing mutations of interest. This can speed up diagnosis and optimize resources since it allows full genome sequencing to be done only on clinically relevant specimens. The assays can be also employed for a rapid and cost-effective detection of VOCs or other variants in sewage for the purposes of wastewater-based epidemiology. The approach proposed here can be used to better understand SARS-CoV-2 variant diversity, geographic distribution and impact worldwide.

## INTRODUCTION

On 31 December 2019, the Wuhan Municipal Health and Health Commission reported a cluster of pneumonia cases of unknown aetiology. Further investigations identified a novel coronavirus as the causative agent of the disease (China CDC, 2020). On 11 March 2020, following the spread of the outbreak to other parts of China and to a number of countries worldwide, the Director General of the WHO declared the COVID-19 outbreak a pandemic (WHO, 2020a). As of 23 February 2021, 111 million cases have been documented in 192 countries, with 2.4 million confirmed deaths (https://coronavirus.jhu.edu/map.html).

A growing number of SARS-CoV-2 variant sequences are being detected since the beginning of the pandemic, some of which considered of global concern. All of these variants have characteristic mutations, the most significant of which are located in the gene encoding the spike (S) protein, potentially affecting viral infectivity and antigenicity (Li et al., 2020; Pillay et al., 2020).

One mutation in the spike protein (D614G), in particular, emerged early in the epidemic and spread rapidly through Europe and North America (Korber et al., 2020; Volz et al., 2020).

In August/September 2020, a SARS-CoV-2 variant linked to infection among farmed minks, subsequently transmitted to humans, was identified in Denmark (Hammer et al., 2020). In all, six countries (Denmark, the Netherlands, Spain, Sweden, Italy and the United States) reported cases of farmed mink infected with SARS-CoV-2 to the World Organisation for Animal Health (OIE) (WHO, 2020b).

An additional novel variant - clade GV, lineage B.1.177 (GISAID) or 20E.EU1 (Nexstrain) - emerged in early summer of 2020 in Spain, and subsequently spread to multiple locations in Europe, possibly introduced into other countries by summer tourists (Hodcroft et al., 2020). The introduction of the B.1.177 lineage into Ontario, Canada, by a traveller returning from Europe in late September 2020 was also reported in a recent preprint (Guthrieet al., 2020).

On 14 December 2020, British authorities announced that a new SARS-CoV-2 variant had been identified through viral genomic sequencing (WHO, 2020c). The variant was initially termed SARS-CoV-2 VUI 202012/01 (Variant Under Investigation, year 2020, month 12, variant 01) and then re-designated VOC (Variant Of Concern) 202012/01 on 18 December 2020 (Chand et al., 2021). The new variant belongs to clade GR, lineage B.1.1.7 (GISAID), or clade 20I/501Y.V1 (Nextstrain) and was discovered following an unforeseen rise in COVID-19 cases in South East England, with a more than three-fold increase in the period from 5 October to 13 December 2020 (WHO, 2020d). As of 19 January 2021, 23 EU/EEA countries, including Italy, have reported the new COVID-19 variant 20I/501Y.V1. In the rest of the world, 37 additional countries reported it (ECDC, 2021b). The emergence of the new SARS-CoV-2 variant, which could be 40-70% more transmissible than previous strains circulating in the UK (WHO 2020b; ECDC, 2020), has prompted some countries to close their borders with the UK. A review by the UK’s New and Emerging Respiratory Virus Threats Advisory Group (NERVTAG) advises that there is moderate confidence that 20I/501Y.V1 has a substantial increase in transmissibility compared to other variants (NERVTAG, 2020; Mahase 2020). However, there is not enough information at present to determine whether this variant is associated with any change in clinical disease severity or increased mortality, though preliminary analyses in the United Kingdom, showed a risk ratio of death of 1.65 for B.1.1.7 cases compared to non-B.1.1.7 cases (PHE, 2021). As for antigenic response, to date, it is considered unlikely that 20I/501Y.V1 may hinder vaccine-induced immunity (Conti et al., 2021). On 18 December 2020, national authorities in South Africa announced the detection of a new variant of SARS-CoV-2, designated VOC-202012/02 (the second Variant Of Concern, December 2020). This variant, belonging to clade GH, lineage B.1.351 (GISAID) or 20H/501Y.V2 (Nextstrain), was reported to be largely replacing other coronavirus strains in South Africa since at least November 2020. As of 19 January 2021, 10 EU/EEA countries (Germany, France, Ireland, Austria, Belgium, Finland, the Netherlands, Norway and Sweden) and 13 countries/territories outside of the EU/EEA have documented the presence of this new SARS-CoV-2 variant (ECDC, 2021b). One case of the South African variant was detected in Italy, in a man who landed at Milan’s Malpensa Airport from Africa at the end of January (https://www.ansa.it/lombardia/notizie/2021/02/03/a-varese-primo-caso-variante-sudafricana-in-italia_41b99448-367c-4000-ac88-9b3ae130d839.html). Preliminary results indicate that it may have increased transmissibility. Here too, as with the VOC 202012/01 variant, there is no evidence that it is associated with a higher severity of infection (ECDC, 2021b).

In January, another variant, clade GR lineage P.1 (GISAID) or 20J/501Y.V3 (Nextrain) was identified in Japan in four travellers who had arrived from Brazil (NIID, 2021; Galloway et al., 2021). Whole genome sequences for all four isolates were deposited in GISAID EpiCoV on 10 January 2021. On 13 January 2021, researchers from Fiocruz Amazônia announced the identification of this variant in the capital of Amazonas, Manaus, where an upsurge in COVID-19 cases was registered in January of 2021 (Farias et al., 2021). To date, the P.1 variant has been identified either in Brazil, or in travellers from Brazil (mostly from the State of Amazonas), including one returning to Italy from Brazil (Maggi et al., 2021).

Lastly, on 17 January 2021, public health officials announced that the SARS-CoV-2 GH, lineage B.1.429 (GISAID) – also termed CAL.20C – has become increasingly common in multiple Californian counties and has been responsible for several large COVID-19 outbreaks in that state. All of these variants have characteristic mutations, the most significant of which are located in the gene encoding the spike (S) protein (Li et al., 2020; Pillay et al., 2020).

The so-called ‘UK variant’ 20I/501Y.V1 is characterized by 23 mutations: 14 nonsynonymous mutations, 3 deletions and 6 synonymous mutations affecting ORF1ab, S, ORF8, and N (Chand et al, 2021). Mutations in the gene that encodes for the S protein, which is associated with viral entry into cells, has attracted the most attention. Ten mutations are documented in the S protein of the UK variant. Two of these, the N501Y (an asparagine to tyrosine amino acid substitution in the receptor binding domain) and the HV69-70del (a 6-base deletion), had already been circulating globally before merging into this new variant. The most relevant changes in the 20I/501Y.V1 variant are likely to be the N501Y mutation and the Y144del - a deletion potentially contributing to spike surface variation. The N501Y mutation is in the receptor binding domain (RDB), important to ACE2 binding and antibody recognition. It is likely to enhance the transmissibility of the virus by improving the S protein’s affinity for the host’s ACE2 receptor. *In vitro* (Starr et al., 2020) and animal experiments (Gu et al., 2020) seem to confirm this hypothesis.

The 20H/501Y.V2 variant, first detected in South Africa, displays the same N501Y mutation (as well as the D614G) as the UK variant, but this mutation is thought to have arisen independently in the two lineages (WHO 2020b). It also carries additional mutations (the most common are L18F, K417N, E484K and K417N) in the S protein, which are not usually present in the UK strain, though mutation E484K has been recently detected in 21 B.1.1.7 sequences in UK (PHE, 2021). Furthermore, 501Y.V2 does not have the HV69-70del characterizing the UK variant, but presents in about 85% of sequences another deletion in the S region, that of amino acids 242 to 244 (LAL242-244del).

The new variant from Brazil, 20J/501Y.V3, harbors 12 mutations in the S protein, some of which it shares with either the ‘UK variant’ (N501Y and D614G) or the ‘South African variant’ (L18F, E484K, N501Y, D614G).

The Spanish variant 20E.EU1 is characterized by substitution A222V in the A domain of the S protein, a domain which is not considered to contribute directly to receptor binding or membrane fusion for SARS-CoV-2 (Hodcroft et al., 2020).

The rapid emergence of variants worldwide highlights the importance of genetic surveillance of the SARS-CoV-2 pandemic. The risk of introduction and community spread of variants of concern in the EU/EEA countries is considered very high (ECDC, 2021b). WHO advises all countries to increase the routine sequencing of SARS-CoV-2 viruses where possible, and to share sequence data internationally through open-source platforms (WHO 2021). ECDC also recommends urgent analysis and sequencing of virus isolates to identify cases of the new variants in a timely manner (ECDC, 2021a). Currently, whole genome sequencing (WGS) of human isolates has resulted in over 591,000 complete genomes available in GISAID as of 23 February, 2021.

A full genome approach (WGS), however, is time-consuming and costly, and requires specialized computational infrastructure and technical skills. Furthermore, WGS cannot be effectively performed in conditions of low virus titres, reduced sequence integrity and simultaneous occurrence of more than one genome.

We developed a long PCR nested RT-PCR assay (∼1500 bp), followed by conventional Sanger sequencing, to quickly screen for the different circulating variants or detect the introduction of possible novel variants. The assay amplifies a key region of the S gene, the region containing the major mutations of the UK, South African and Brazilian variants, as well as crucial mutations of other variants of clinical interest, like the 20E.EU1 that emerged in Spain, the mink-associated variant, and the CAL.20C variant identified in California (see Table 1). Preliminary studies on sequences available in GISAID and Nextclade were performed before application on field samples. The first, aimed at verifying whether specific mutation combinations are sufficiently informative to screen for specific SARS-CoV-2 variants, and the second, a comparative study of full-length sequences and their corresponding ∼1500 bp fragments to ascertain whether the different variants would be correctly assigned to their respective clades on the sole basis of the genome region amplified by the long nested PCR assay.

**Table 1.**
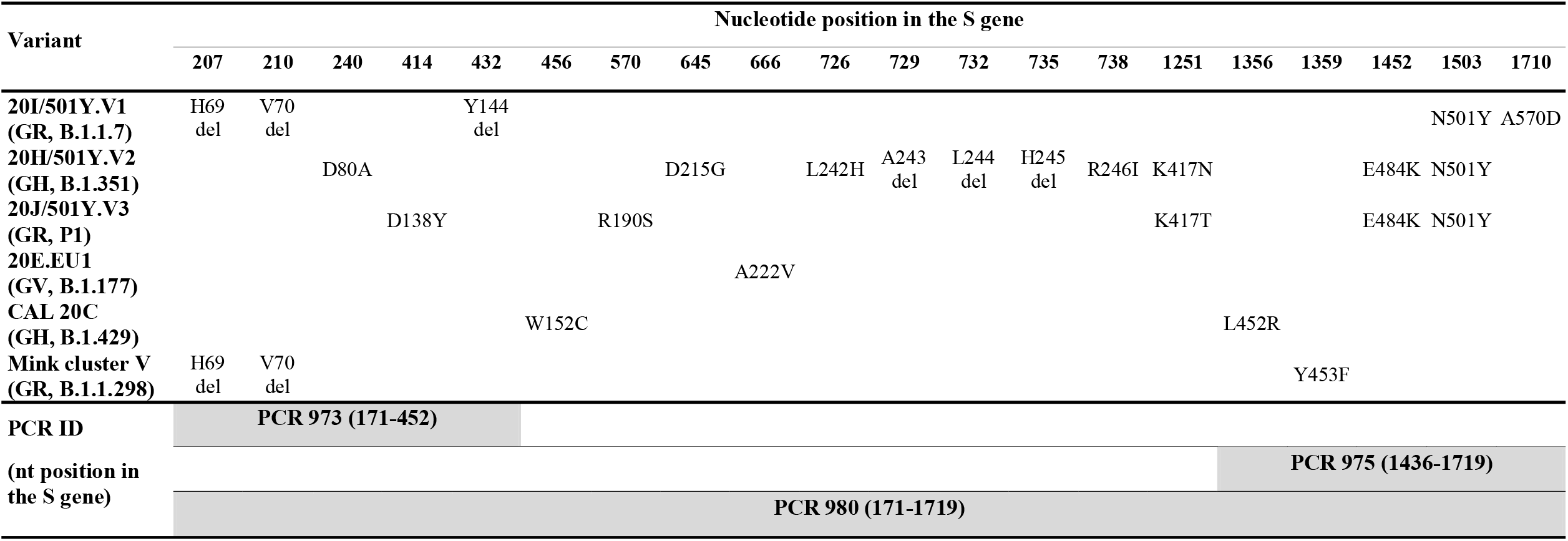
Mutations indicative of the main SARS-CoV-2 VOC detectable by the newly designed PCR assays.

We subsequently enhanced our method to allow variant detection in highly challenging matrices such as wastewater, where the amplification of long RNA fragments is difficult, by designing two additional short nested RT-PCRs targeting portions of the region spanned by the long assay.

SARS-CoV-2 is excreted by both symptomatic and asymptomatic individuals that are infected with the novel coronavirus, and ends up in wastewater systems (Foladori et al., 2020). By analysing wastewater, it is therefore possible to obtain information on viruses circulating in the population served by a given Wastewater Treatment Plant (WTP). Previous studies have detected SARS-CoV-2 RNA in wastewater samples worldwide, highlighting environmental surveillance as a valuable tool potentially allowing us to detect peaks in virus circulation and estimate the proportion of individuals infected with SARS-CoV-2 in a given catchment area (Hart et al., 2020), thus contributing to inform public health interventions.

All the assays were successfully tested on a 20I/501Y.V1 strain detected in Italy at the end of December 2020 (see Supplementary material), and on a Wuhan strain (clade 19A). Finally, the three assays were tested on a panel of field samples of clinical (viral isolates from swabs) and environmental (sewage) origin.

The methods proposed could be used to perform targeted sequencing of clinical and environmental samples for screening and early detection of SARS-CoV-2 variants.

## MATERIALS AND METHODS

### Primer design and preliminary in silico studies

Primers were designed to capture key mutations of different variants. Such mutations (Table 1), potentially affecting viral infectivity and antigenicity, are located in the gene encoding the spike (S) protein. Primers (Table 2 and Figure 1) were designed using the Primer 3 software (http://www.bioinformatics.nl/cgi-bin/primer3plus/primer3plus.cgi).

**Table 2:**
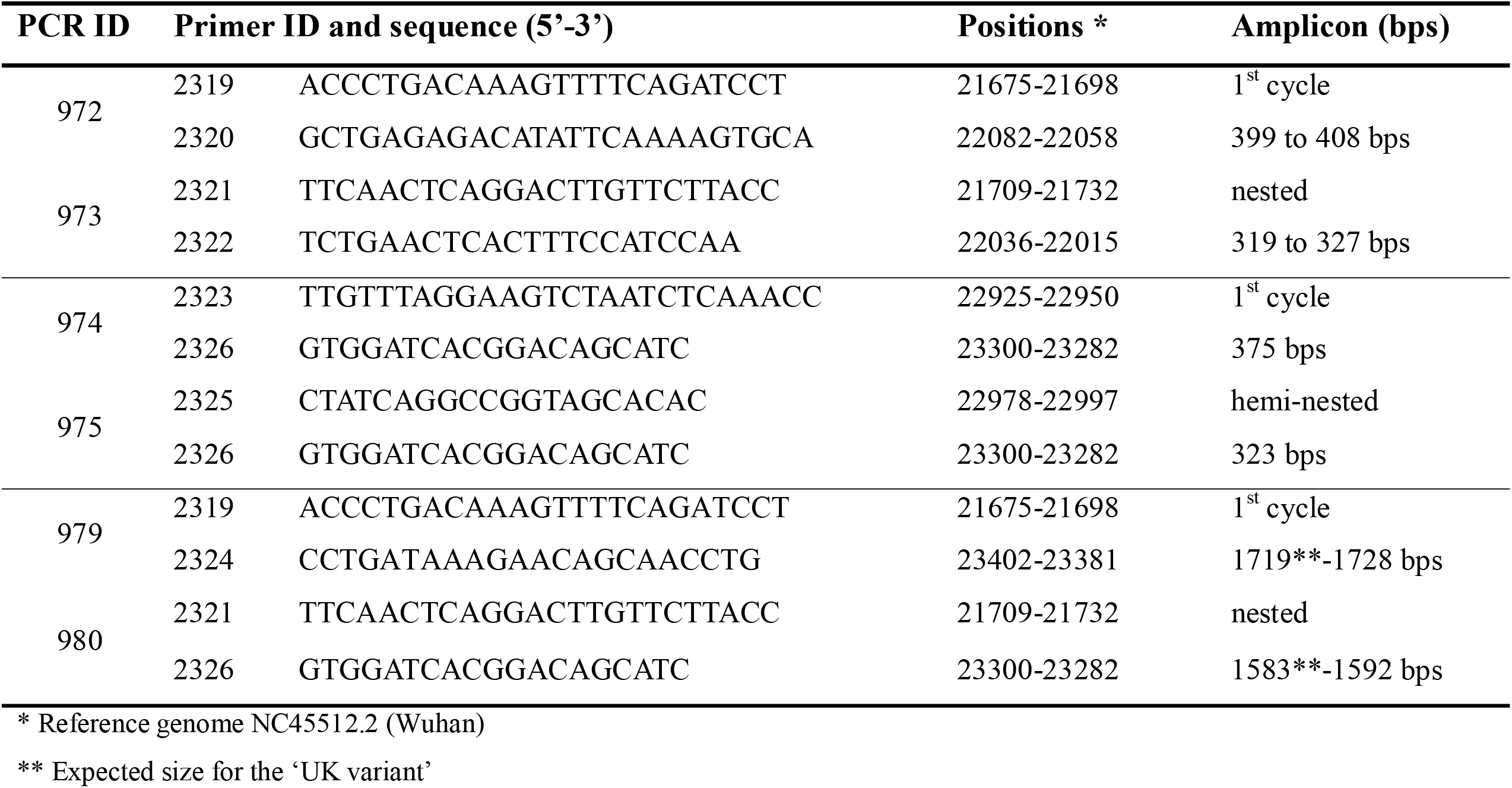
Primers and PCRs used in this study.

**Figure 1.**
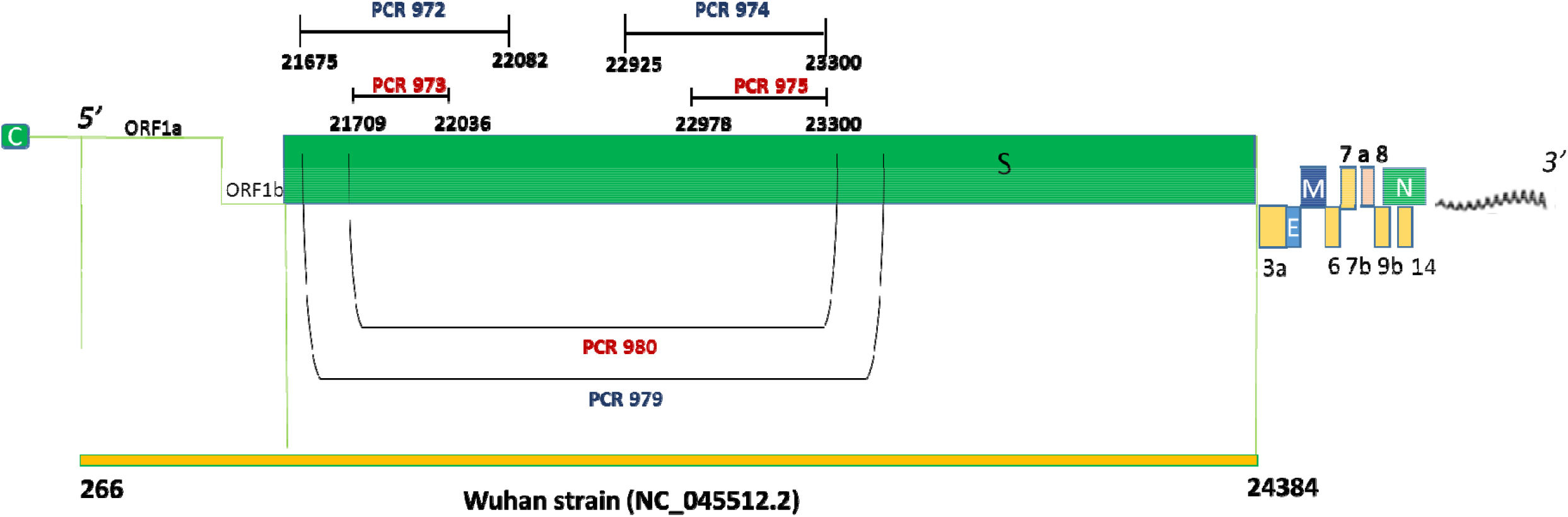
Positions of the primers in the newly designed PCR assays in the Spike gene of SARS-CoV-2.

Ultimately, three assays were designed with different aims:

1. A long nested RT-PCR (first cycle - PCR ID 979; nested assay - PCR ID 980, 1583-1592 bps) designed to screen for the presence of mutations characteristic of the three VOCs (20I/501Y.V1, 20H/501Y.V2, and 20J/501Y.V3), as well as other variants of interest such as 20E.EU1,CAL 20C, and mink variant, as shown in Table 1.
2. A short nested RT-PCR (319-327 bps) amplifying a portion of the region targeted by the long assay (first cycle - PCR ID 972; nested assay - PCR ID 973) designed to reveal the presence of the mutations HV69-70del and Y144del (UK variant), D80A (South African variant), and D138Y (Brazilian variant).
3. Another short nested RT-PCR (323 bps) amplifying a different portion of the region targeted by the long assay (first cycle - PCR ID 974; hemi-nested assay - PCR ID 975) designed to reveal the presence of the key mutation N501Y shared by different variants of concern. This amplicon can also reveal the presence of amino acid substitution A570D (combined with N501Y) of the 20I/501Y.V1 variant, and E484K (combined with the N501Y) of either the 20H/501Y.V2 or the 20J/501Y.V3 variant. It can also detect L452R of the Californian variant and Y453F of the mink variant.

Before application to field samples, a preliminary in silico study was performed to examine the discriminatory potential of the specific mutations covered by our long assay, for SARS-CoV-2 variant identification. This was done using GISAID (https://www.gisaid.org/) and Nextclade (https://nextstrain.org/). The GISAID study was performed on 20I/501Y.V1 (14 January 2021) and on 20H/501Y.V2 (17 January 2021) using complete genomes (see Supplementary Materials). Overall, considering the characteristic mutations of 20I/501Y.V1 present in the ∼ 1500 bp fragment of the S gene chosen for the long assay (HV69-70del, Y144del, N501Y and A570D), the results showed that among the complete genomes belonging to lineage GR B.1.1.7 any combination of two mutations was present at a frequency ranging from 97.6% to 99.4%. Furthermore, cluster analysis attributed all of the sequences displaying at least two of the aforementioned mutations to the GR B.1.1.7 lineage. As for the 20H/501Y.V2 variant, characteristic mutations such as D80A, E484K, N501Y, LAL242-244del were present at a frequency ranging from 88.6 to 91.4%. Any combination of two of these mutations was present in clade GH lineage B.1.351 at a frequency exceeding 84.2%, and sequences displaying at least two of the mutations belonged to lineage B.1.351 in >99% of cases. One significant exception was the E484K/N501Y combination, which belonged to lineage GH B.1.351 in 96.8% of cases, due to its presence in the 20J/501Y.V3 (Brazilian) variant as well. Based on this preliminary analysis, it was concluded that, for these two variants, the detection of a combination of at least two mutations in the selected ∼ 1500 bp fragment provided a strong indication of the presence of the variant in question.

To further explore the discriminatory potential of our newly-designed long nested PCR for the different SARS-CoV-2 variants, we compared the ∼1500 bp fragments to their corresponding full-length sequences in terms of how they would be classified by Nextclade web tool. We therefore created two FASTA files: one containing 30 full-length genomes, comprising five GISAID strains for each of the following clades/variants: 19A (Wuhan, China), 20I/501Y.V1 (UK), 20H/501Y.V2 (South Africa), 20J/501Y.V3 (Brazil), 20E.EU1 (Spain), CAL.20C (California) as described in the Supplementary Materials; and the other, derived from the first, containing the corresponding ∼1500 bp fragments amplified by our long nested PCR assay. Both files were then analyzed using the Nextclade web tool (nextstrain.org) in order to determine whether our nested assay (PCR ID 980) would yield similar results to those given by a full sequence analysis. Nextclade web tool assigned all the variants correctly on the sole basis of the genome region amplified by the PCR ID 980. The only significant exception was the 20I/501Y.V1 variant, which was incorrectly assigned to clade 20B. This misclassification, however, was due to a limitation of the Nextclade web tool, which, for the purposes of phylogenetic placement and clade assignment, uses only mutations and no gaps (https://github.com/nextstrain/nextclade/issues/311#issuecomment-765904855), and was therefore unable to take into consideration the deletions HV69-70del and Y144del for a correct assignment of our partial sequences of the S gene. Similarly, the two trees built with Nextclade, one based on full-length genomes and the other based on ∼1500 bp fragments, both correctly assigned all of the variants to the branches corresponding to clades 19A, 20H/501Y.V2, 20J/501Y.V3 and 20E.EU1. Only the ‘UK variant’ was placed at the base of the 20I/501Y.V1 branch (Figure 2).

**Figure 2.**
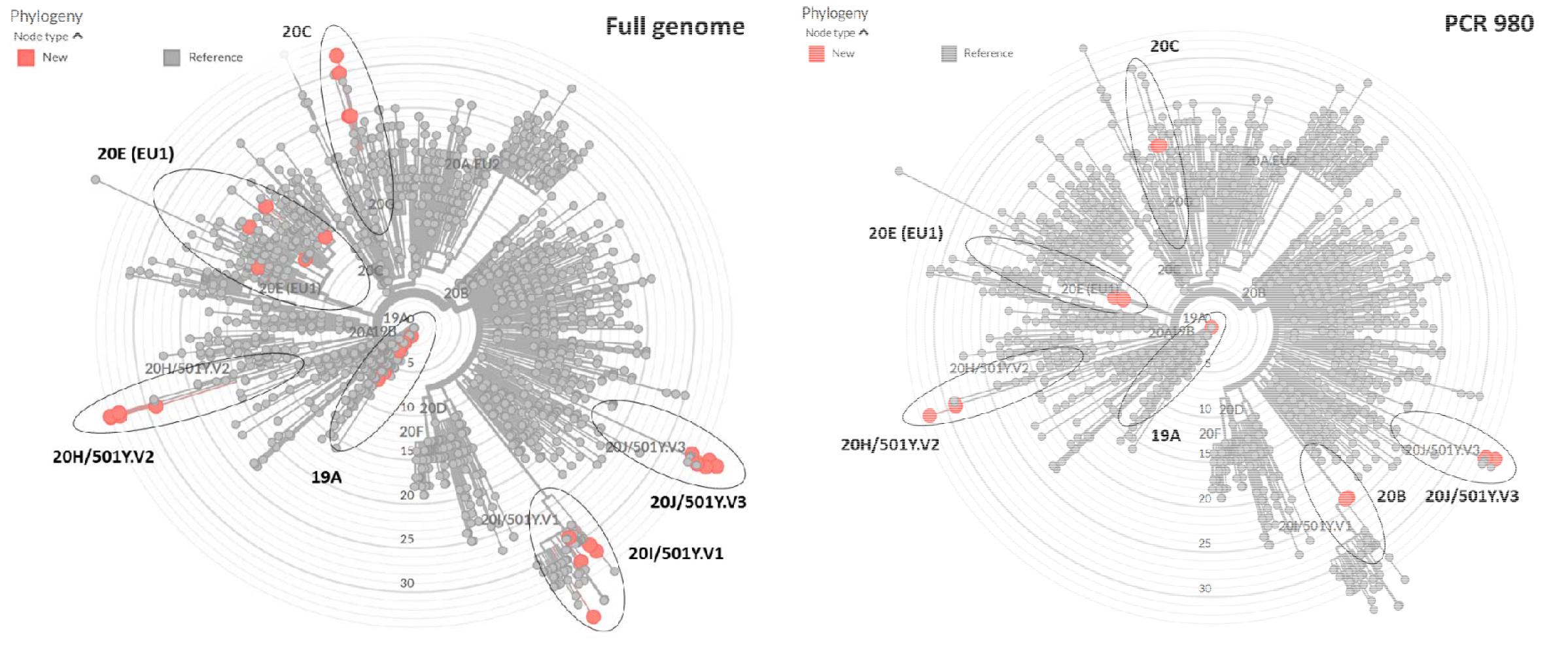
Nextclade web tool phylogenetic trees for representative strains of the different variants using the full genome or the Spike partial gene fragment amplified with PCR ID 980 - nextstrain.org with ad hoc modifications.

### Primer validation on field samples and assay specificity

The RNA of the 20I/501Y.V1 variant was obtained from a nasopharyngeal swab of an Apulian patient who had returned from the United Kingdom. The sample had previously been isolated on Vero E6 cells, fully sequenced (Experimental Zooprophylactic Institute of Puglia and Basilicata), and the sequence submitted to GISAID under reference hCoV-19/Italy/APU-IZSPB-399PT/2020. RNA from the Wuhan strain (kindly provided by the Robert Koch-Institute under the RefBio project) was included as control.

Before being tested with the three nested PCR assays, both RNAs were standardized at a concentration ranging between 10^2^ and 10^3^ genome copies (g.c.)/μL using a previously published real-time RT-qPCR (La Rosa et al., 2021).

The European Virus Archive global (EVAg) Coronavirus RNA specificity panel (kindly provided by the Erasmus University Medical Center, Rotterdam, The Netherlands), which includes both Alfa- and Beta-coronaviruses (HCoV-NL63, HCoV-229E, HCoV-OC43, MERS-CoV, SARS-CoV and SARS-CoV-2), was used to assess the specificity of our newly-designed primers.

### Field samples

#### i) Clinical samples

Seven SARS-CoV-2 RNA samples had previously been extracted from viruses originating from nasopharyngeal swabs following isolation on Vero E6 cells, and had been characterized by WGS and deposited in GISAID as belonging to clades 20A, 20B, 20E.EU1 (two samples each), and 20I/501Y.V1 (one sample). Viral concentrations in these samples were calculated as previously described (La Rosa et al., 2021) and found to be in excess of 10^6^ g.c./μL. The samples were subsequently diluted to an approximate concentration of 10^4^ g.c./μL before testing by nested PCR.

#### ii) Environmental samples

a total of 34 sewage samples were included in the study. Twenty samples, collected in Wastewater Treatment Plants (WTPs) in Rome, Italy, between September and December 2020 (see Supplementary Materials), had been previously analyzed and found positive for SARS-CoV-2 following the procedure described in La Rosa et al. (2021). Briefly, concentration of sewage samples (250 ml) was performed by the two-phase polyethylene glycol-dextran separation method (WHO 2003) and nucleic acids were extracted using the NucliSens extraction system (bioMerieux, Marcy-l’Etoile, France). The eluted RNA (100 µl) was aliquoted and stored at –80°C until molecular analysis. Viral concentrations in these samples ranged from 2.0 to 37.8 g.c./μL of RNA (corresponding to 1.6×10^3^ to 3.0×10^4^ g.c./L of raw sewage).

Eight sewage samples (4 composite and 4 grab) were collected between 21 and 25 January 2021 in 4 WTPs in the small town of Guardiagrele in Abruzzo (central Italy), where a cluster linked to the UK variant had been reported, to investigate whether the S mutations associated with 20I/501Y.V1 were detectable in the local sewage.

Furthermore, six sewage samples were collected between 5 and 8 February 2021 in the city of Perugia, Umbria (central Italy), where the UK variant had been on the rise and the Brazilian variant had been identified as well (https://www.regione.umbria.it/documents/18/25027633/Ordinanza+n.14+del+6+febbraio+2021.pdf).

### Nested RT-PCR conditions

Following standardization of the protocol, SuperScript IV One-Step RT-PCR System with Platinum SuperFi RT-PCR Master mix (Invitrogen, Carlsbad, CA, USA) was used for the RT-PCR. The amplification conditions for the RT-PCR were as follows: reverse transcription at 45 °C for 10 min, denaturation at 98 °C for 2 min, followed by 35 cycles of 98 °C for 10 sec, 58 °C for 10 sec, 72°C for either 30 sec (PCR ID 972 and 974) or 1 min (PCR ID 979), and a final extension at 72 °C for 5 min. Each of the three PCR reactions was performed using 1 μL of each primer (10 μM; Table 1) and either 4 μL of RNA in a final volume of 25 μL (clinical samples) or 8 μL of RNA in a final volume of 50 μL (environmental samples). After the first round of PCR, nested PCR was performed using the Phusion Hot Start II DNA Polymerase with GC buffer (Thermo Fisher Scientific, Waltham, MA, USA), 2 μL of the first PCR product, 1 μL of each primer (10 μM) and the following conditions: initial denaturation at 98 °C for 30 sec, followed by 45 cycles of 98 °C for 10 sec, 62 °C for 30 sec, 72 °C for either 30 sec (PCR ID 973 and 975) or 1 min (PCR ID 980), and a final extension at 72 °C for 10 min. Standard precautions were taken to avoid laboratory contamination. PCR products were observed by gel electrophoresis (2% agarose gel, stained with GelRed, Biotium; Fremont, CA, USA). To confirm amplifications, all products were purified from the PCR reaction or from gel using either Montage PCRm96 Micro-well Filter Plate (Millipore, Burlington, MA, USA) or GRS PCR & Gel Band Purification Kit (GRISP, Porto, Portugal) and were sequenced (see Supplementary Materials) on both strands (Bio-Fab Research, Rome, Italy and Eurofins Genomics, Ebersberg, Germany).

Sequences were submitted to GenBank under the accession numbers [to be assigned; submission IDs 4852387, 4852507, and 4852527].

### Bioinformatics analysis

The public database GISAID (Shu and McCauley, 2017) was used for BLAST searches and for mutation analysis. The Nextclade web tool v.012.0 (Hadfield et al., 2018) was used to compare study sequences to SARS-CoV-2 reference sequences, assign them to clades, and determine their position within the SARS-CoV-2 phylogenetic tree. As of 21 January 2021, the database includes 11 major clades (19A, 19B, 20A, 20B, 20C, 20D, 20E(EU1), 20F, 20G, 20I/501Y.V1, 20H/501Y.V2, 20J/501Y-V3). Auspice v2.0, an open-source interactive tool, was used to visualize phylogenetic trees.

All SARS-CoV-2 mutations found in field samples were compared with the GISAID reference strain hCoV-19/Wuhan/WIV04/2019 using the CoVsurver mutation analysis tool (https://www.gisaid.org/epiflu-applications/covsurver-mutations-app/), which provides geographic and temporal distributions of SARS-CoV-2 mutations.

## RESULTS

### Primer validation and assay specificity

All three newly designed PCR assays successfully amplified the two strains (the UK variant and the Wuhan strain) used for primer assessment.

Amplicon sequencing of the long fragment of the UK variant revealed the presence of mutations HV69-70del, Y144del, N501Y and A570D characteristic of 20I/501Y.V1 (Table 1). Sequencing of the short amplicons obtained using the other two assays confirmed this result. In the Nextclade tree constructed using the ∼1500 bps sequence, this isolate fell at the base of the branch corresponding to clade 20I/501Y.V1, as predicted by the preliminary study we conducted with the UK variant in Nextrain prior to primer design (Figure 2). As for the Wuhan strain, although amplification was achieved with all three assays, the long PCR yielded a low amount of product, probably due to RNA fragmentation, allowing only partial sequencing of the fragment. Identity with GenBank reference sequence NC_045512 was confirmed for amplicons provided by the two short assays.

Specificity of the three designed assays was confirmed on the EVAg Coronavirus RNA panel. Amplicons of the expected size were obtained only for SARS-CoV-2 (sequencing confirmed 100% identity to reference sequence NC_045512) while no amplification was present for HCoV-NL63, HCoV-229E, HCoV-OC43, MERS-CoV, and SARS-CoV RNAs.

### Clinical samples

All of the seven SARS-CoV-2 RNAs extracted from viruses cultured on Vero E6 cells and previously characterized by WGS were successfully amplified by the three newly developed assays. Sequencing results of amplicons generated by the long PCR are summarized in Table 3. The geographic and temporal occurrence of the mutations detected in the field samples, obtained using the GISAID CoVsurver: Mutation Analysis tool, is summarized in Supplementary Materials.

**Table 3:** Variant analysis of clinical samples obtained using PCR ID 980 and different bioinformatics tools.

| Sample ID | Identification based on WGS | Mutation map <sup>1</sup> | GISAID blast | Nextclade |
| --- | --- | --- | --- | --- |
| swab_25 | G B.1 / 20A | - | Not assignable <sup>2</sup> | 19A |
| swab_26 | G B.1 / 20A | D215H | Not assignable <sup>3</sup> | 19A |
| swab_27 | GR B.1.1.316 / 20B | - | Not assignable <sup>2</sup> | 19A |
| swab_28 | GR B.1.1.229 / 20B | - | Not assignable <sup>2</sup> | 19A |
| swab_29 | GV B.1.177 / 20E EU1 | A222V | GV_B.1.177 | 20E (EU1) |
| swab_30 | GV B.1.177 / 20E EU1 | A222V | GV_B.1.177 | 20E (EU1) |
| swab_31 | GR B.1.1.7 / 20I/501Y.V1 | H69del, V70del, Y144del, N501Y, A570D | GR/501Y.V1 (B.1.1.7) | 20B |

<sup>1</sup> Analysis with GISAID CoVsurver: Mutation Analysis (https://www.gisaid.org/epiflu-applications/covsurver-mutations-app/)
<sup>2</sup> 100% identity with sequences assigned to the following clades: GR\_B.1.1.222; GR\_B.1.1.70; GR\_B.1.1.119; GR\_B.1.1.28; GR\_B.1.1.316; GR\_B.1.1.33; GR\_B.1.1.103; GH\_B.1.2; GH\_B.1.425; G\_B.1; G\_B.1.91; GH\_B.1.4703
<sup>3</sup> 100% identity with sequences assigned to the following clades: G\_B.1; GH\_B.1.2; GR\_B.1.1.47; GR\_B.1.1.304; GR\_B.1.1.127; GR\_B.1.1.284; GR\_B.1.1.119; O\_B-13
n.d. not done due to absence of PCR amplification

Sequences obtained by the long PCR confirmed the presence of the two variants in swab samples (20E.EU1 in swab IDs 29 and 30, and 20I/501Y.V1 in swab ID 31). The UK variant, however, could only be assigned to its correct lineage using the GISAID BLAST tool, given the aforementioned limitation of the Nextclade tool. Samples free of mutations or carrying mutations not specific to any VOC in the analyzed region (swab IDs 25 to 28) could not be differentiated and assigned. Indeed, they showed 100% nt identity with different clades/lineages in GISAID, and were all classified as 19A by Nextclade (Table 3).

All clinical samples also tested positive using the two short nested PCR assays. Amplicon sequencing of the short PCR fragments confirmed the sequences obtained using the long nested PCR in all samples.

### Environmental samples

A total of 34 wastewater samples were tested with the three nested PCR assays. PCR and sequencing results are summarized in Table 4. The long nested RT-PCR did not amplify any of the 20 SARS-CoV-2-positive urban sewage samples collected from WTPs in Rome (IDs 3702 to 3815). Amplification, however, was achieved with PCR IDs 973 and 975 (8 and 10 samples, respectively, with an overlap of 5 samples). None of these samples displayed mutations indicative of VOCs.

**Table 4.** Variant analysis of environmental samples obtained using PCR IDs 980, 973, and 975.

| Sample ID | Sampling location | PCR ID 980 |  | PCR ID 973 |  | PCR ID 975 |  |
| --- | --- | --- | --- | --- | --- | --- | --- |
|  |  | result | mutations | result | mutations | result | mutations |
| Archive samples |  |  |  |  |  |  |  |
| 3716 | Rome East | - |  | - |  | + | none |
| 3732 | Rome North | - |  | + | none | + | none |
| 3737 | Rome East | - |  | - |  | + | none |
| 3738 | Rome East | - |  | + | none | + | none |
| 3740 | Rome East | - |  | - |  | + | none |
| 3742 | Rome Ostia | - |  | + | none | + | none |
| 3750 | Rome North | - |  | - |  | + | none |
| 3798 | Rome East | - |  | + | none | + | none |
| 3801 | Rome East | - |  | + | none | - |  |
| 3802 | Rome East | - |  | - |  | + | none |
| 3805 | Rome East | - |  | + | none | - |  |
| 3808 | Rome South | - |  | + | none | - |  |
| 3809 | Rome Ostia | - |  | + | none | + | none |
| Ad-hoc samples from outbreak locations |  |  |  |  |  |  |  |
| 3862 | Guardiagrele | - |  | + | none | + | none |
| 3863 | Guardiagrele | + | A222V | + | none | + | none |
| 3865 | Guardiagrele | + | A222V | + | none | - |  |
| 3944 | Perugia | - |  | + | D138Y | + | E484K, N501Y |
| 3945 | Perugia | - |  | + | D138Y | + | E484K, N501Y |
| 3947 | Perugia | + | D138Y, R190S, K417T, E484K, and N501Y | + | D138Y | + | E484K, N501Y |
| 3949 | Perugia | - |  | + | E96G | + | N501Y, A570D |

Samples IDs 3702, 3714, 3720, 3739, 3744, 3797, 3815, 3864, 3868, 3869, 3946 and 3948 did not provide amplification in any of the assays and were not included in the table.

With regard to the 14 sewage samples collected from locations experiencing outbreaks of SARS-CoV-2 variants, the long nested PCR successfully amplified two of the eight samples collected in Guardiagrele, Abruzzo (IDs 3863 and 3865) and one of the six samples collected in Perugia, Umbria (ID 3947). The first two samples showed mutations characteristic of the Spanish variant (A222V), while the sample ID 3947 carried the mutation characteristic of the Brazilian variant (D138Y, R190S, K417T, E484K, and N501Y). The short assays - PCR ID 973, and PCR ID 975 - successfully amplified 7 and 6 of the 14 sewage samples, respectively (with an overlap of 6 samples, including the three samples testing positive also by the long assay).

Sequencing of fragments obtained by PCR 973 showed no mutations in the PCR amplicons originating from Guardiagrele. On the contrary, this short assay did find the presence of mutation D138Y, characteristic of the Brazilian variant, in three of the six samples collected in Perugia: ID 3944, 3945, and 3947. Another sample from Perugia (ID 3949), showed the aa substitution E96G, while the remaining two amplifications did not display any mutation.

The second short assay, PCR 975, confirmed the results obtained by PCR 973 on the amplified samples from Guardiagrele (IDs 3862 and 3863), namely, the absence of mutations indicative of VOCs. In the four samples collected in Perugia and identified as carrying mutations, on the other hand, this short assay (PCR 975) showed the key mutation N501Y. This mutation was combined with E484K in three of them (IDs 3944, 3945, and 3947), a combination present in both the Brazilian and the South African variants (see Table 1). The results of PCR 973, where all three samples showed the aa substitution D138Y, and those of the long assay showing one of them (ID 3847) to carry all 5 aa substitutions typical of the Brazilian variant, pointed to the presence of this VOC. The fourth sample from Perugia (ID3949) carried mutation N501Y along with A570D, a key signature of the UK variant.

## DISCUSSION

SARS-CoV-2 sequence diversity was initially thought to be low, but an increasing number of variant sequences have since been detected, indicating that new mutations are continually emerging. Major variants discovered thus far include the UK variant (clade GR, lineage B.1.1.7), the South African variant (GH, lineage B.1.351), the Brazilian variant (clade GR, lineage P1), the variant from California (GH, B.1.429), the Spanish variant, the mink variant, and others. All these SARS-CoV-2 variants carry characteristic mutations (deletions and/or missense mutations) in different parts of the genome, some in important regions in terms of potential clinical significance, such as the gene coding for the spike protein (the S gene). The spike protein appears to be critical for cellular entry, and allows the virus to attach to the host cell (Pillar et al., 2020), which is why this protein is also a target for vaccine development and antigen testing.

The analysis of SARS-CoV-2 virus genomes can complement and support strategies to reduce the burden of COVID-19 (WHO, 2021). Whole genome sequencing has proven to be a powerful tool capable of helping us understand outbreak transmission dynamics and spill-over events and to screen for mutations with a potential impact on transmissibility, pathogenicity or countermeasures (e.g. diagnostics, antiviral drugs and vaccines) (WHO, 2020d). For this reason, the WHO recommends that routine sequencing of SARS-CoV-2 viruses be increased where possible, and sequence data be shared through open-source platforms (WHO 2020c). The ECDC similarly encourages EU/EEA states to sequence at least 500 randomly selected samples each week, and to prioritize samples from large outbreaks, or samples collected from travellers and individuals with an infection detected 2 weeks after vaccination (ECDC, 2021b).

Whole genome sequencing demands substantial investment in terms of time and financial resources, however, and the acquisition of sufficient, high quality RNA to maximize sequencing yield and the validity of sequence data. In addition, bioinformatic analysis necessitates high-performance computational power and trained staff. To address some of these problems, we developed a rapid, inexpensive and sensitive long nested PCR assay to detect key changes in the S protein of SARS-CoV-2, indicative of the presence of VOCs 20I/501Y.V1, 20H/501Y.V2 and 20J/501Y.V3, as well as of other variants of clinical interest. This assay can be used as a first screening tool to select samples of interest for WGS characterization. Resources for genome sequencing would thus be optimized by allocating them to clinically relevant specimens. The region amplified by this long assay was specially chosen to reveal key mutations distinctive of main variants, as shown in Table 1.

The discriminatory potential of the long PCR assay was initially explored in silico using GISAID and Nextclade. Phylogenetic trees predicted the ∼1500 bps sequence of the S protein to have the same discriminatory power as the entire genome in assigning sequences with the S mutations characteristic of the VOCs to their corresponding clusters. The ability of primer pairs to amplify SARS-CoV-2 RNA and the discriminatory power of the amplified fragments were then tested on field samples, consisting of SARS-CoV-2 viral isolates, already been fully sequenced. All of the clinical samples were successfully amplified with the long PCR. The 20I/501Y.V1 and 20E.EU1 variants present in the panel were correctly identified by the long assay. Other variants of concern, like the Brazilian or the South African variants, were not available to us as field samples. Indeed, only one case of the South African variant had been detected in Italy, in a man who landed at Milan’s Malpensa Airport from Africa at the end of January (https://www.ansa.it/lombardia/notizie/2021/02/03/a-varese-primo-caso-variante-sudafricana-in-italia_41b99448-367c-4000-ac88-9b3ae130d839.html). Having downloaded the sequence (GISAID EPI_ISL_1012924), we verified that it harbors the Spike mutations D80A, D215G, L242del, L244del, K417N, E484K, and N501Y - all of which would have been detectable by our long PCR assay. Similarly, only three cases of the Brazilian variant had been identified and fully sequenced (GISAID EPI_ISL_875566 to ID EPI_ISL_875568) thus far in Italy. All these isolates carry the characteristic Spike mutations D138Y, R190S E484K, K417T, and N501Y included in our assay. Once we had confirmed that the developed PCR assay could be successfully used to screen for variants in clinical samples, we went on to investigate whether this assay could also be used for the screening of SARS-CoV-2 variants in complex matrices, such as urban wastewaters. Since the amplification of long fragments poses specific challenges in such matrices, we also designed two short nested RT-PCR assays targeting portions of the region spanned by the long assay, to be used in case of long PCR failure. A number of studies have demonstrated the added value of regular surveillance of wastewaters in combination with other indicators for the management of the pandemic and a recent document by the European Commission highlights the importance of wastewater surveillance to allow screening among large population groups (European Commission, 2021). This can help public health authorities identify where more detailed analysis is needed and speed up the detection of variants as part of an increased genomic and epidemiological surveillance. Two recent papers have demonstrated that environmental surveillance can be used to study SARS-CoV-2 diversity (Crits-Christophet al, 2021; Martin et al., 2020). Crits-Christoph and co-workers sequenced SARS-CoV-2 RNA directly from sewage collected in the San Francisco Bay Area to generate complete and near-complete SARS-CoV-2 genomes, and found that the major consensus genotypes detected in the sewage were identical to clinical genomes from the region. Moreover, additional variants not found in clinical samples, were identified in wastewaters, suggesting that wastewater sequencing can provide evidence for recent introductions of viral lineages before they are detected by local clinical sequencing (Crits-Christophet al, 2021). Metagenomic analysis of environmental matrices is a complex task, however, with high operational costs and specific requirements in terms of bioinformatic skills.

Martin and co-workers demonstrated changes in virus variant predominance during the COVID-19 pandemic by tracking SARS-CoV-2 in sewage using a nested RT-PCR approach targeting five different regions of the viral genome (Martin et al., 2020). Here, we adopted a similar approach, but concentrated on a unique region of the SARS-CoV-2 genome - the spike protein gene, which harbors several key mutations associated with known VOCs and other variants of interest.

The newly-designed assays were therefore used on 34 sewage samples, including 20 SARS-CoV-2-positive samples collected from WTPs in Rome between September and December 2020, eight sewage sample collected from a small town in Abruzzo where an outbreak of the UK variant was reported in mid-January (https://www.ansa.it/abruzzo/notizie/2021/01/18/covid-a-guardiagrele-36-casi-da-variante-inglese_fbfc1c88-649b-4f32-9042-d1f0648b2bbb.html), and six sewage samples collected in Perugia, Umbria, where, at the beginning of February, both UK and Brazilian variants have been documented by public health authorities (https://www.regione.umbria.it/documents/18/25027633/Ordinanza+n.14+del+6+febbraio+2021.pdf).

As expected, only three of the samples could be amplified and successfully sequenced with the long PCR assay, one from Umbria, the others from Abruzzo. Wastewater is indeed a complex matrix where the amplification of long fragments is seldom achieved, due to the presence of inhibitors, nucleic acids of different origin, and fragmented target genomes, often in low amounts. The sequences obtained by the long assay from samples collected in January in Abruzzo showed mutations characteristic of the Spanish variant, which was indeed the most common variant in the EU till December 2020 (ECDC, 2021a). The sequence obtained from Perugia displayed instead the mutations indicative of 20J/501Y.V3. Interestingly, this sample was collected on 6 February 2021 from a WTP serving the outskirts of Perugia, including the local hospital, which had reportedly been experiencing nosocomial clusters of this variant, as per press releases issued by public health officials (https://www.ansa.it/umbria/notizie/2021/02/05/rezza-in-umbria-variante-brasiliana-in-cluster-ospedalieri_d50f6dce-9b78-443c-a0d8-0d23e16058a8.html; https://tg24.sky.it/salute-e-benessere/2021/02/08/covid-umbria).

As expected, the two short nested RT-PCR assays were more sensitive than the long one, successfully amplifying about half of the tested sewage samples. No mutations of interest were detected in the samples collected in Rome and in Guardiagrele, Abruzzo, using the short assays, as the mutation A222V, characteristic of the Spanish variant, is not included in the short fragments. Conversely, sequences obtained from the sewage samples collected in Perugia, Umbria, showed the presence of mutations characteristic of the UK variant (one sample) and of the Brazilian variant (three samples, one of which was confirmed by the long assay). Both of these variants had been circulating in the area according to statements by public health authorities, but no genome sequences were available at the time of the study.

Comparing results obtained by the two nested PCRs for the same sample, concordant results were obtained for the Brazilian variant (presence of mutation D138Y in the fragment obtained by PCR 973, and mutations E484K/N501Y in the amplicon of PCR 975). In contrast, the sample harboring mutations of the UK variant in the region amplified by PCR 975 (N501Y and A570D) did not show the expected mutations (HV69-70del and Y144del) in the region amplified by PCR 973. This exposes a limit of the combined use of two short PCR assays. Since different SARS-CoV-2 strains may coexist in a single sewage sample, the use of separate tests may result in each of the tests amplifying a sequence belonging to a different strain. However, our GISAID in silico study showed the combination of the N501Y and A570D mutations to strongly suggest that the sequence carrying them belongs to lineage GR B.1.1.7. Furthermore, we only performed classical Sanger sequencing on PCR amplicons, and this approach may underestimate the existence of some, possibly less common, sequences. Our next goal is therefore to combine the protocol with NGS on PCR amplicons for a more in-depth analysis of sequences, as successfully applied to enteric viruses (Suffredini et al., 2018; Iaconelli et al, 2017).

To our knowledge, this is the first evidence of the presence of sequences harboring key mutations of 20I/501Y.V1 and 20J/501Y.V3, as well as of the Spanish variant in urban wastewaters, highlighting the potential contribution of wastewater surveillance to explore SARS-CoV-2 diversity. Moreover, we demonstrated the potential of this approach, based on nested amplification followed by conventional Sanger sequencing, to allow the rapid identification of specific VOCs in the catchment of any given WTP to quickly identify areas where the intensification of clinical surveillance and/or targeted preventive intervention is required.

In conclusion, the method we propose can be used to rapidly screen SARS-CoV-2-positive samples for VOCs and other variants of clinical interest. Compared with WGS, conventional RT-PCR followed by Sanger sequencing can be performed routinely at low cost and does not require specialized expertise to interpret the results. Through the targeting of key mutations, this approach could enable the rapid screening of samples and the identification of samples to submit to WGS. Moreover, the long nested RT-PCR described here can provide significant, albeit partial, molecular information for samples in which low amounts or poor quality SARS-CoV-2 RNA hamper the successful application of WGS. The approach can also be applied to environmental samples for the purposes of SARS-CoV-2 wastewater-based epidemiology, to explore the possible presence of VOCs or other variants of clinical interest, therefore highlighting geographical areas where an enhancement of clinical monitoring could be beneficial.

Due to an increase in the prevalence of new SARS-CoV2 variants with possible clinical implications, monitoring the spread of different variants in the general population in a timely, cost-effective manner is of utmost importance. The proposed method can contribute to achieve this goal.

## Data Availability

Sequences have been deposited in GenBank

## Acknowledgments

This publication was supported by the European Virus Archive Global (EVA-GLOBAL) project, which provided the coronavirus RNA panel for the assessment of our in-house PCR. The EVA-GLOBAL project, in turn, received funding from the European Union’s Horizon 2020 research and innovation programme under grant agreement No 871029.

Our thanks go to “Project RefBio - Germany’s contribution to strengthen the bio-analytical reference laboratories in the UNSGM (2017-2021), EQAE on SARS-CoV-2 (2020), Centre for Biological Threats and Special Pathogens (ZBS 1-3) at RKI (Project Coordinator), Berlin, Germany” for providing the Wuhan strain.

We also thank Stefano Fontana for his insightful comments.

We thank Luigina Serrecchia, Viviana Manzulli, Valeria Rondinone, Ines della Rovere, Fiorenza Petruzzi and Mr. Guido Asturio for their technical support.

We acknowledge the support of Utilitalia, Italian federation of energy, water and environmental services, and ACEA ATO2 for wastewater sampling in Rome. We also thank Tommaso Pagliani and Fabio Campitelli of SASI SpA (Società Abruzzese per il Servizio Idrico Integrato) and the Municipal Authorities for allowing wastewater collection in Guardiagrele (CH), Abruzzo, and Tiziana Buonfiglio, Andrea Vitali, Paolo Meniconi,Massimo Lorenzetti Renzetti and Marino Burini, Umbra Acque SpA, for allowing wastewater collection in Perugia, Umbria.

