## supplementary for "Rapid Screening for SARS-CoV-2 Variants of Concern in Clinical and Environmental Samples Using Nested RT-PCR Assays Targeting Key Mutations of the Spike Protein"

**Study in GISAID on the predictive capacity of specific mutations for the identification of SARS-CoV-2 variants**

**VOC 202012/01 (“UK variant”) - GISAID 20I/501Y.V1 - clade GR, lineage B.1.1.7**

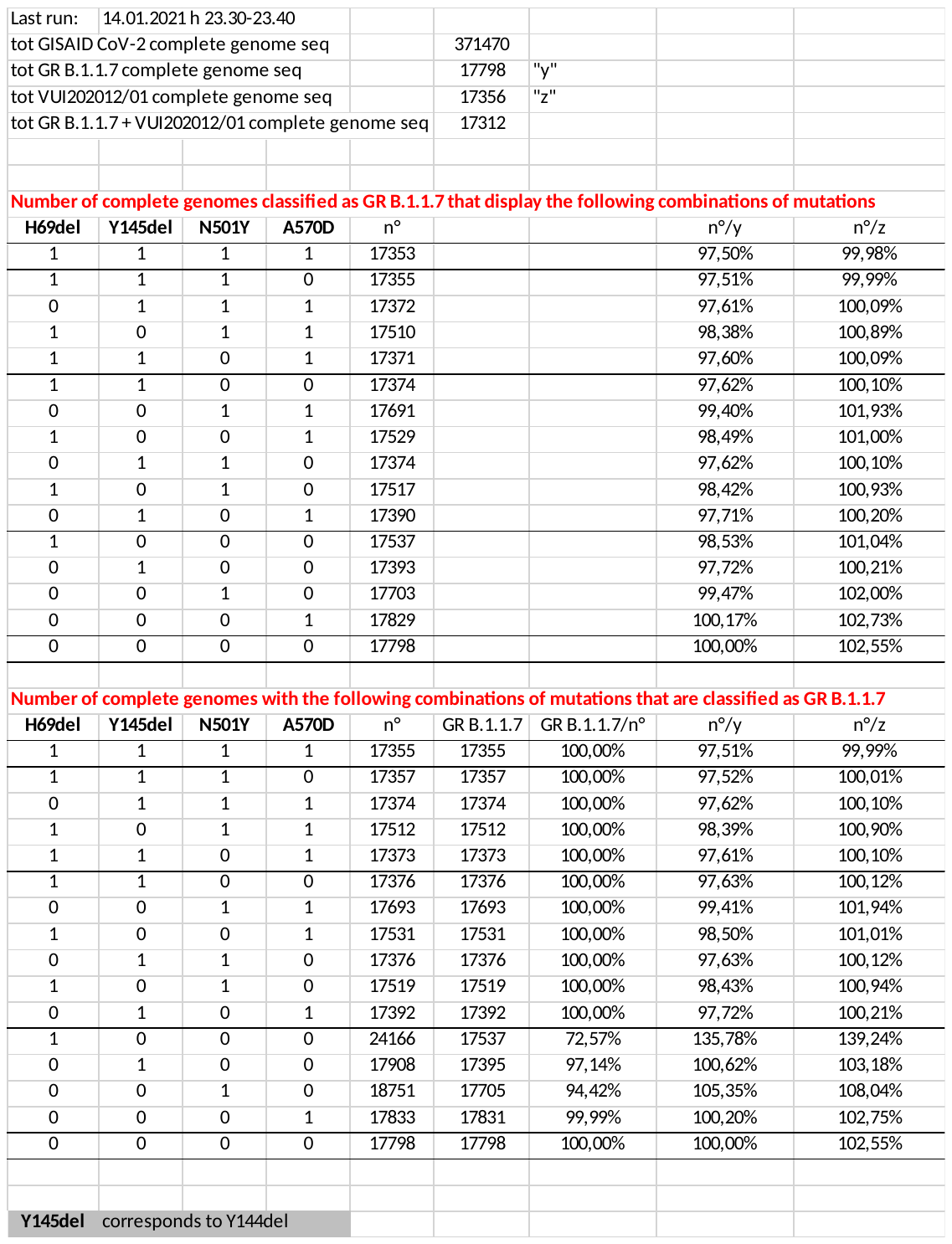

**VOC 202012/02 (“South Africa”) - GISAID 20H/501Y.V2 - clade GR, lineage B.1.351**

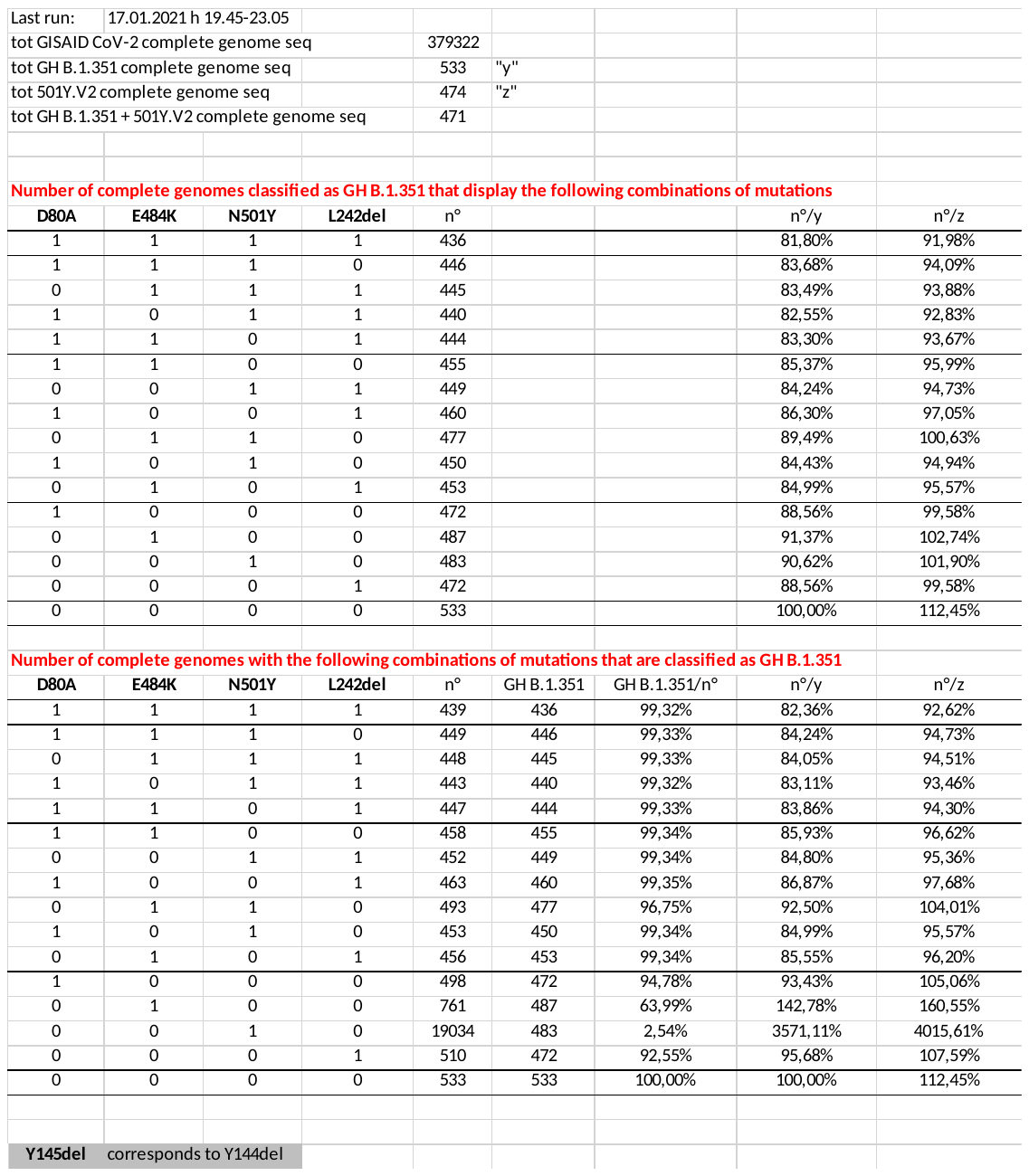

**Confirmation of the discriminatory capacity of the PCR ID 980 in Nextclade (https://clades.nextstrain.org/)**

Sequences (full genome or partial Spike gene sequence included in PCR ID 980) used for the study

| **Clade** | **GISAID sequence ID** | **Nextclade** | |
| --- | --- | --- | --- |
|  |  | **Full genome** | **PCR 980** |
| 20I/501Y.V1  (UK variant) | hCoV-19/Denmark/DCGC-23017/2020\|EPI ISL 793399\|2020-12-28 | 20I/501Y.V1 | 20B |
|  | hCoV-19/Portugal/PT2159/2021\|EPI ISL 801520\|2021-01-02 |  |  |
|  | hCoV-19/Norway/7072/2020\|EPI ISL 813977\|2020-12-21 |  |  |
|  | hCoV-19/Netherlands/NH-RIVM-21371/2020\|EPI ISL 826536\|2020-12-28 |  |  |
|  | hCoV-19/England/205390794/2021\|EPI ISL 846595\|2021-01-02 |  |  |
| 20H/501Y.V2  (South African variant) | hCoV-19/South Africa/KRISP-EC-K005341/2020\|EPI ISL 678626\|2020-11-12 | 20H/501Y.V2 | 20H/501Y.V2 |
|  | hCoV-19/South Africa/NHLS-UCT-GS-2090-KRISP/2020\|EPI ISL 696452\|2020-11-19 |  |  |
|  | hCoV-19/South Africa/NHLS-UCT-GS-3558-KRISP/2020\|EPI ISL 696459\|2020-11-20 |  |  |
|  | hCoV-19/South Africa/NHLS-UCT-GS-7043-KRISP/2020\|EPI ISL 696494\|2020-11-24 |  |  |
|  | hCoV-19/SouthAfrica/KRISP-K007135/2020\|EPI ISL 825130\|2020-11-24 |  |  |
| 20J/501Y.V3 (Brazilian variant) | hCoV-19/Brazil/AM-20843269RC/2020\|EPI ISL 833140\|2020-12-23 | 20J/501Y.V3 | 20J/501Y.V3 |
|  | hCoV-19/Japan/IC-0562/2021\|EPI ISL 792681\|2021-01-02 |  |  |
|  | hCoV-19/Brazil/AM-994/2020\|EPI ISL 833174\|2020-12-22 |  |  |
|  | hCoV-19/Japan/TY7-501/2021\|EPI ISL 833366\|2021-01 |  |  |
|  | hCoV-19/South Korea/KDCA0464/2021\|EPI ISL 833249\|2021-01-10 |  |  |
| 20E.EU1  (Spanish variant) | hCoV-19/Ireland/WW-NVRL-AIIDV1206v1/2020\|EPI ISL 768773\|2020-09-25 | 20E (EU1) | 20E (EU1) |
|  | hCoV-19/Spain/CT-IBV-97012007/2020\|EPI ISL 855493\|2020-09-18 |  |  |
|  | hCoV-19/Italy/CAM-COTUGNO-11204/2020\|EPI ISL 855573\|2020-12-21 |  |  |
|  | hCoV-19/Denmark/DCGC-32260/2020\|EPI ISL 856676\|2020-12-07 |  |  |
|  | hCoV-19/Belgium/ULG-11309/2021\|EPI ISL 856753\|2021-01-13 |  |  |
| CAL 20C  (variant common in California) | hCoV-19/USA/NM-CDC-2-3768096/2021\|EPI_ISL_903839\|2021-01-02 | 20C | 20C |
|  | hCoV-19/USA/CT-CDC-2-3774026/2020\|EPI_ISL_903921\|2020-12-03 |  |  |
|  | hCoV-19/USA/CA-CDC-2-3774040/2020\|EPI_ISL_903892\|2020-12-31 |  |  |
|  | hCoV-19/USA/CA-LACPHL-AF00262/2021\|EPI_ISL_905771\|2021-01-10 |  |  |
|  | hCoV-19/USA/WA-OHSU-9371/2021\|EPI_ISL_906034\|2021-01-07 |  |  |
| 19A  (Wuhan strain) | hCoV-19/Italy/FVG-ISS-2249/2020\|EPI ISL 856893\|2020-03-01 | 19A | 19A |
|  | hCoV-19/Ireland/D-20G41967/2020\|EPI ISL 848165\|2020-03-26 |  |  |
|  | hCoV-19/USA/NY-AECOM 075/2020\|EPI ISL 826638\|2020-04-07 |  |  |
|  | hCoV-19/Italy/LAZ-INMI-SPL1/2020\|EPI ISL 412974\|2020-01-29 |  |  |
|  | hCoV-19/Italy/LOM-UniMI-L228/2020\|EPI ISL 542215\|2020-03-11 |  |  |

**Field samples details**

***Clinical samples***

| **Clinical sample ID** | **Viral**  **isolation ID** | **Sampling date** | **GeneFinderTM Covid-19 Plus RealAmp Kit** | | **La Rosa et al., 2021**  **nsp14 region** | |
| --- | --- | --- | --- | --- | --- | --- |
|  |  |  | **RdRp gene** | **N gene** |  |  |
|  |  |  | **Ct** | **Ct** | **Ct** | **g.c./μL** |
| swab_25 | 407 | dec-2020 | 15,6 | 15,6 | 13,17 | 2,21E+07 |
| swab_26 | 414 | dec-2020 | 16,5 | 15,8 | 14,11 | 1,18E+07 |
| swab_27 | 170 | aug-2020 | 14,4 | 14,4 | 10,85 | 1,03E+08 |
| swab_28 | 172 | aug-2020 | 16,1 | 17,0 | 15,03 | 6,37E+06 |
| swab_29 | 413 | dec-2020 | 14,3 | 14,4 | 10,46 | 1,34E+08 |
| swab_30 | 423 | dec-2020 | 14,6 | 14,6 | 11,02 | 9,24E+07 |
| swab_31 | 399 | dec-2020 | 16,2 | 14,9 | 13,10 | 1,74E+07 |

| **Clinical sample ID** | **GISAID sequence ID** | **GISAID identification**  **based on WGS** | **Nextstrain identification**  **based on WGS** |
| --- | --- | --- | --- |
| swab_25 | hCoV-19/Italy/APU-IZSPB 407PT/2020\|EPI ISL 794749\|2020-11-25 | G B.1 | 20A |
| swab_26 | hCoV-19/Italy/APU-IZSPB 414PT/2020\|EPI ISL 794755\|2020-11-25 | G B.1 | 20A |
| swab_27 | hCoV-19/Italy/APU-IZSPB-170APT/2020\|EPI ISL 649190\|2020-08-17 | GR B.1.1.316 | 20B |
| swab_28 | hCoV-19/Italy/APU-IZSPB-172APT/2020\|EPI ISL 649191\|2020-08-20 | GR B.1.1.229 | 20B |
| swab_29 | hCoV-19/Italy/APU-IZSPB 413PT/2020\|EPI ISL 794754\|2020-12-16 | GV B.1.177 | 20E EU1 |
| swab_30 | hCoV-19/Italy/APU-IZSPB 423PT/2020\|EPI ISL 794759\|2020-12-22 | GV B.1.177 | 20E EU1 |
| swab_31 | hCoV-19/Italy/APU-IZSPB-399PT/2020\|EPI ISL 745193\|2020-12-21 | GR B.1.1.7 | 20I/501Y.V1 |

***Environmental samples***

| **Sewage sample ID** | **WWTP ID** | **Sampling date** |
| --- | --- | --- |
| ***Archive samples*** | | |
| 3702 | Rome Ostia | 26/09/2020 |
| 3714 | Rome East | 05/10/2020 |
| 3716 | Rome East | 14/10/2020 |
| 3720 | Rome South | 12/10/2020 |
| 3732 | Rome North | 26/10/2020 |
| 3737 | Rome East | 04/11/2020 |
| 3738 | Rome East | 04/11/2020 |
| 3739 | Rome East | 13/11/2020 |
| 3740 | Rome East | 13/11/2020 |
| 3742 | Rome Ostia | 11/11/2020 |
| 3744 | Rome East | 19/11/2020 |
| 3750 | Rome North | 11/11/2020 |
| 3797 | Rome East | 17/11/2020 |
| 3798 | Rome East | 17/11/2020 |
| 3801 | Rome East | 02/12/2020 |
| 3802 | Rome East | 02/12/2020 |
| 3805 | Rome East | 14/12/2020 |
| 3808 | Rome South | 12/12/2020 |
| 3809 | Rome Ostia | 11/12/2020 |
| 3815 | Rome South | 26/12/2020 |
| ***Ad-hoc samples from outbreak locations*** | | |
| 3862 | Guardiagrele (CH), Abruzzo | 21/01/2021 |
| 3863 | Guardiagrele (CH), Abruzzo | 26/01/2021 |
| 3864 | Guardiagrele (CH), Abruzzo | 26/01/2021 |
| 3865 | Guardiagrele (CH), Abruzzo | 26/01/2021 |
| 3866 | Guardiagrele (CH), Abruzzo | 26/01/2021 |
| 3867 | Guardiagrele (CH), Abruzzo | 26/01/2021 |
| 3868 | Guardiagrele (CH), Abruzzo | 26/01/2021 |
| 3869 | Guardiagrele (CH), Abruzzo | 26/01/2021 |
| 3944 | Perugia, Umbria | 05/02/2021 |
| 3945 | Perugia, Umbria | 05/02/2021 |
| 3946 | Perugia, Umbria | 06/02/2021 |
| 3947 | Perugia, Umbria | 06/02/2021 |
| 3948 | Perugia, Umbria | 08/02/2021 |
| 3949 | Perugia, Umbria | 08/02/2021 |

**Primer used for sequencing the PCR fragment of PCR ID 980 (long nested assay)**

| **Primer ID** | **Sequence (5’-3’)** | **Positions *** | **Strand** |
| --- | --- | --- | --- |
| 2321 | TTCAACTCAGGACTTGTTCTTACC | 21709-21732 | forward |
| 2322RevCom | TTGGATGGAAAGTGAGTTCAGA | 22036-22015 | forward |
| 2323 | TTGTTTAGGAAGTCTAATCTCAAACC | 22925-22950 | forward |
| 2320 | GCTGAGAGACATATTCAAAAGTGCA | 22082-22058 | reverse |
| 2325RevCom | GTGTGCTACCGGCCTGATAG | 22978-22997 | reverse |
| 2326 | GTGGATCACGGACAGCATC | 23300-23282 | reverse |

* Reference genome NC45512.2 (Wuhan)

**Geographic and temporal occurrence of variants detected in clinical samples using PCR ID 980**

| **Sample ID** | **Mutation map** | **Mutation occurrence**  **(GISAID CoVsurver Mutation Analysis)** |
| --- | --- | --- |
| swab_29, swab_30 | A222V | AA change Spike A222V already occurred 93796 times (23.50% of all samples with Spike sequence) in 58 countries. The first strain with this aa change, collected in February 2020, was hCoV-19/Iran/K1r-29/2020. The aa change most recently occurred in strain hCoV-19/England/SHEF-10CF750/2021, collected in January 2021. |
| swab_25, swab_27, swab_28 | - | /// |
| swab_25 | Swab 25 provided a double electropherogram signal, one of which including a nine amino acid deletion:  IHVSGTNGT 68-76del | N74del and T76del. Mutation Spike N74del and T76del removes a potential N-glycosylation site at position 74, which may also affect antigenic and other properties of this strain. In detail, the motif at positions 74-76 changed from NGT (glyco) to --- (no glyco). |
| swab_26 | [D215H](javascript:Spike_D215H();) | AA change Spike D215H already occurred 1155 times (0.32% of all samples with Spike sequence) in 22 countries. The first strain with this aa change, collected in February 2020, was hCoV-19/Hangzhou/ZJU-010/2020. The aa change most recently occurred in strain hCoV-19/USA/NY-Wadsworth-21002030-01/2021, collected in January 2021. |
| swab_31 | [H69del](javascript:Spike_H69del();), [V70del](javascript:Spike_V70del();),  Y144del, N501Y,  A570D | AA change Spike N501Y already occurred 19062 times (5.23% of all samples with Spike sequence) in 45 countries. The first strain with this aa change, collected in April 2020, was hCoV-19/Brazil/PE-IAM19/2020. The aa change most recently occurred in strain hCoV-19/England/210260926/2021, collected in January 2021.  AA change Spike A570D already occurred 18023 times (4.95% of all samples with Spike sequence) in 42 countries. The first strain with this aa change, collected in April 2020, was hCoV-19/Italy/ABR-IZSGC-TE26533/2020. The aa change most recently occurred in strain hCoV-19/Singapore/48/2021, collected in January 2021. |
